# Estimate of the rate of unreported COVID-19 cases during the first outbreak in Rio de Janeiro

**DOI:** 10.1101/2021.10.08.21264741

**Authors:** M.S. Aronna, R. Guglielmi, L.M. Moschen

## Abstract

In this work we fit an epidemiological model SEIAQR (*Susceptible* - *Exposed* - *Infectious* - *Asymptomatic* - *Quarantined* - *Removed*) to the data of the first COVID-19 outbreak in Rio de Janeiro, Brazil. Particular emphasis is given to the unreported rate, that is, the proportion of infected individuals that is not detected by the health system. The evaluation of the parameters of the model is based on a combination of error-weighted least squares method and appropriate B-splines. The structural and practical identifiability is analyzed to support the feasibility and robustness of the parameters’ estimation. We use the bootstrap method to quantify the uncertainty of the estimates. For the outbreak of March-July 2020 in Rio de Janeiro, we estimate about 90% of unreported cases, with a 95% confidence interval (85%, 93%).

## 1. Introduction

In late December 2019, health professionals in the city of Wuhan (Hubei, China) identified several cases of pneumonia (The 2019 nCoV Outbreak Joint Field Epidemiology Investigation Team and Q. Li, 2020) caused by a new coronavirus, which was named SARS-CoV-2. The disease induced by SARS-CoV-2, called COVID-19, rapidly spread around the world. Most COVID-19 cases are asymptomatic patients or with mild symptoms, but in more severe cases the disease may progress to viral pneumonia and multi-organ failure (World Health Organization, 2020) and can lead to hospitalization and death.

Brazil declared the disease as a public health emergency in February 2020, before the first COVID-19 cases were reported in Brazil. With the aim of protecting the population, several measures were implemented, including the adjustment of a legal framework to carry out isolation and quarantine (Croda et al., 2020). In March 2020, the governor of the state of Rio de Janeiro declared a public health emergency and ordered to avoid gatherings, closing down schools, restaurants, and theaters, and restricting access to beaches, shopping centers, and non-essential commerce (Dantas et al., 2020). Nevertheless, these measures could not prevent the outbreak of the virus in the region, fueled by the large portion of asymptomatic individuals and by the long incubation period of the virus (Li et al., 2020). The objective of this work is to provide a quantitative estimate of crucial parameters related to the COVID-19 outbreak in the city of Rio de Janeiro during the period March-July 2020. The analysis is based on the fitting of the SEIAQR compartmental model introduced in (Aronna et al., 2021) to real data retrieved from the public health agency (Municipal Health Department, City Hall of Rio de Janeiro, 2021). This epidemiological model takes into account isolation, quarantine of confirmed cases, and testing of asymptomatic individuals as non-pharmaceutical strategies to contain the spread of the virus among the population (more details in Section 2.1). The main purpose of the study is to estimate the rate of unreported cases in the city of Rio de Janeiro. This is a crucial step to gauge the real extent and impact of the disease in the population, since most of the infections do not result in severe symptoms and are therefore likely to remain undetected.

The text is organized as follows: Section 2 collects different information regarding the mathematical model; the structure of the data as retrieved for the city of Rio de Janeiro; and the representation chosen for the parameters’ estimation. Section 3 describes the notions of identifiability analyzed in the paper. Section 4 presents the results of the fitting of the data to the model and the resulting estimates of the parameters. Finally, the last two sections conclude the paper with some discussions and final comments.

### 1.1. Data Availability

No new data are released as part of this research. This paper relies on publicly available datasets, with references provided in the text. The code required to reproduce our analysis is available in the GitHub repository (Moschen, 2021).

## 2. Material and methods

### 2.1. The epidemiological model

We recall the key features of the compartmental model from (Aronna et al., 2021). The population is split into the compartments *S, E, I, A, Q, R*, and *D*, corresponding to the compartments of susceptible, exposed, infectious, infectious and asymptomatic, quarantined or hospitalized, recovered, and dead individuals, respectively. These compartments are linked in the following way (see Figure 1): the individuals in *S* move to compartment *E* when exposed to the virus. After a given latent period, they become infectious and thus pass to compartment *I*. At this stage, an individual in *I* may either report symptoms and thus move (after testing) to the compartment *Q*, or result in an asymptomatic/paucisymptomatic infection and move to the compartment *A*. Individuals from *E* and *A* may move to *Q* as a result of the testing strategy among the asymptomatic population, as described in more detail in the following. Finally, the compartment *R* collects the recovered individuals from either *A* or *Q*, whereas the compartment *D* counts the COVID-19-related deaths. By normalizing the total population to 1, the value of each variable *S, E, I, A, Q, R*, and *D* represents the proportion of that given compartment in the total population. Moreover, we neglect the birth and natural death rates, given the limited time horizon chosen for the data fitting. The dynamics is described by the following set of differential equations:

**Figure 1:**
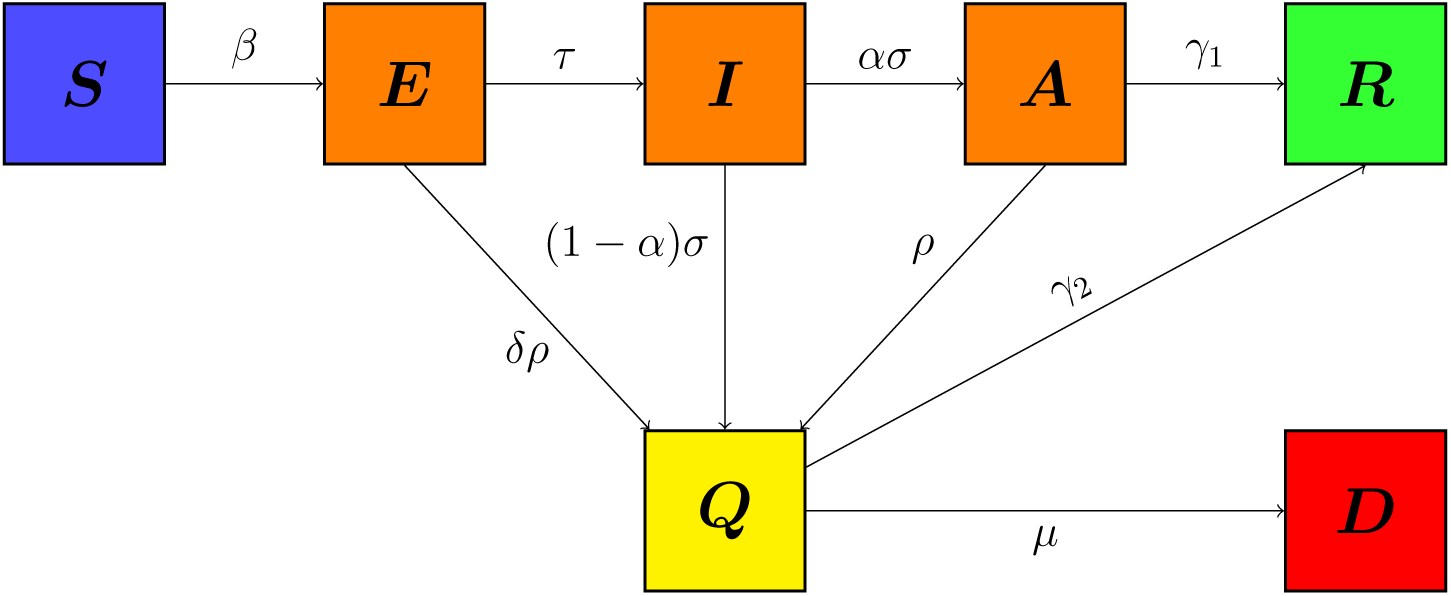
Diagram of the compartmental model

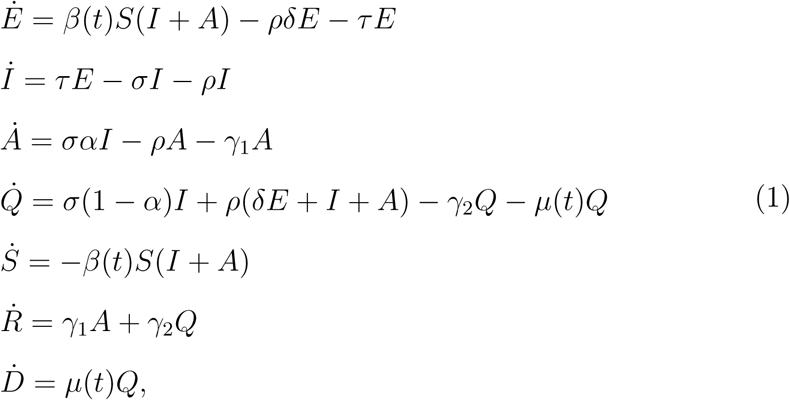

which is a simplified version of the model studied in (Aronna et al., 2021). More precisely, the simplification is done by considering the population as a sole homogeneous group that is subject, in average, to the same mobility restrictions, while the model from (Aronna et al., 2021) allowed for a distinction between two groups - one with high mobility consisting of essential workers and the other with restricted mobility composed by the remainder of the population. The parameters *τ, σ, ω, γ*_1_, *γ*_2_, and *µ* related to the pathogen and the induced disease are described in Table 1. The function *β*(*t*) is the *effective contact rate* at time *t*, which takes into account the average contact rate - directly affected by social distancing, public policies, use of Personal Protective Equipment (PPE), etc. - and the transmissibility of the virus - probability of infection given a contact between an infected and a susceptible individual. A constant of particular interest in our analysis is the parameter *α* ∈ (0, 1), which represents the proportion of asymptomatic infectious cases. Among these cases, only those found positive through either testing induced by contact tracing or simply random testing are reported to the health system. All the others will remain unreported, being paucisymptomatic (but infectious) cases. In view of the scarcity of testing kits during the first outbreak in Rio de Janeiro, we consider the parameter *α* as a proxy for the *unreported rate* of infections. It is clear that estimating such parameter *α* is crucial to assess the effective size of the epidemic since such unreported cases may fuel the ongoing outbreak. Moreover, having an approximation of the size of the recovered population is also an essential information for epidemiological management and the comprehension of the virus behaviour. At the same time, estimating *α* may be very difficult, since it accounts for those cases not detected by the radar of the health system (Nogrady, 2020).

**Table 1:**
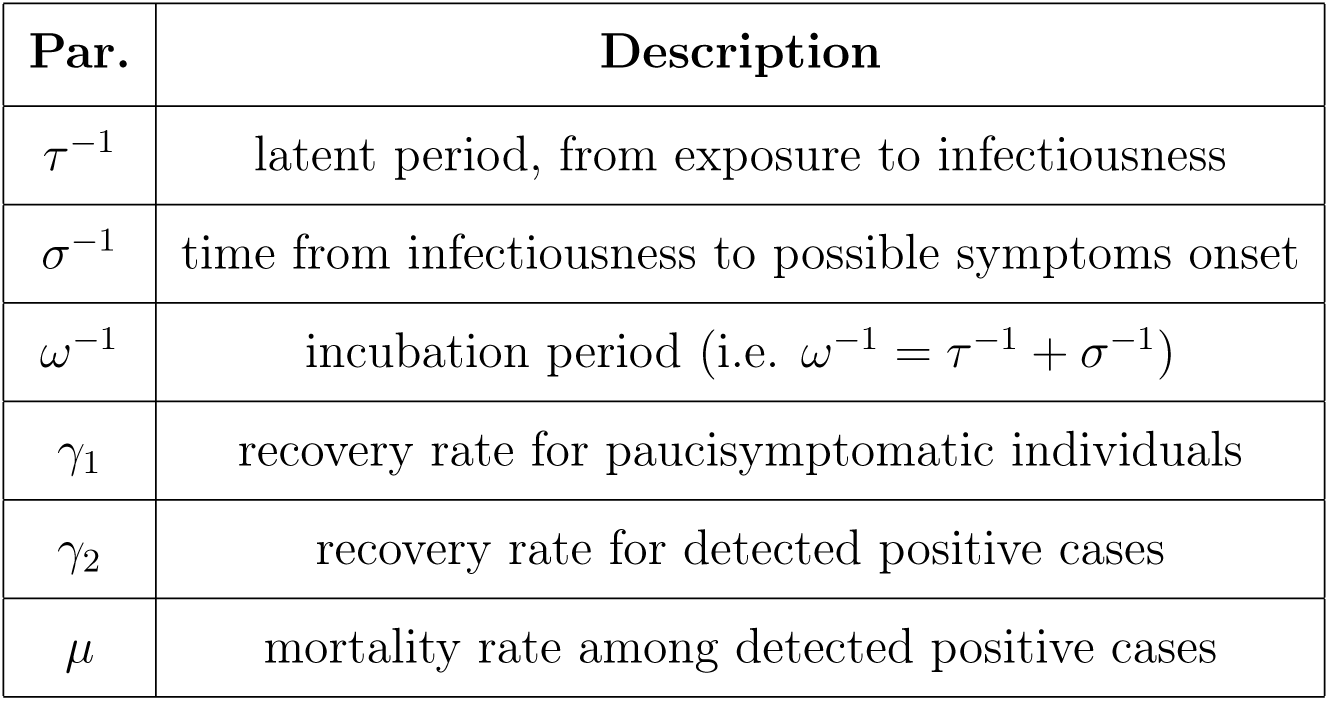
Parameters of COVID-19

| Par. | Description |
| --- | --- |
| $\tau^{-1}$ | latent period, from exposure to infectiousness |
| $\sigma^{-1}$ | time from infectiousness to possible symptoms onset |
| $\omega^{-1}$ | incubation period (i.e. $\omega^{-1} = \tau^{-1} + \sigma^{-1}$) |
| $\gamma_1$ | recovery rate for paucisymptomatic individuals |
| $\gamma_2$ | recovery rate for detected positive cases |
| $\mu$ | mortality rate among detected positive cases |

As mentioned above, the unreported rate is strictly related to the availability of testing kits and to the effectiveness of tracing, tracking, and testing in place during the outbreak. Such measures are described in model (1) by both the detection rate 1 − *α* and the parameter *ρ*, the latter representing the *rate of testing among asymptomatic or paucisymptomatic individuals*. We point out that the value of the unreported rate *α* is not sensitive to small variations of values of *ρ* (see Section 2.3.1).

For clarity of exposition, in model (1) we do not describe important features such as the *sensitivity* and the *specificity* of the testing kits, although these attributes may play a crucial role in consideration of new variants of the virus (see (Aronna et al., 2021, Remark 2.3) for more details). With this premise, in our model we assume that a person in the compartments *I* or *A* will always test positive, in *S* always negative, and in *E* positive with a probability *δ* ∈ (0, 1). We also introduce the counter *T* (*t*) of total positive tests, which follows the dynamics

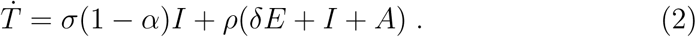

#### 2.1.1. The basic reproduction number

Assuming a constant contact rate *β*, the basic reproduction number ℛ_0_ associated with the model (1) is given by the relation

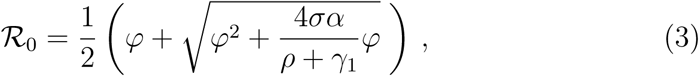

where

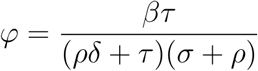

(see (Aronna et al., 2021)). However, as the epidemic evolves, the recovered and immune portion of the population becomes more relevant, impacting the force of the spread. For this reason we introduce the effective (time-dependent) reproduction number ℛ_*t*_, which decreases as the susceptible population *S*(*t*) decreases and it is expressed by the same relation (3) of ℛ_0_ with *φ* replaced by

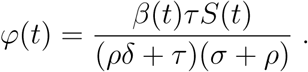

### 2.2. Data description

We retrieve data from the public health agency (Municipal Health Department, City Hall of Rio de Janeiro, 2021), collecting information on COVID-19 confirmed cases and deaths in the municipality of Rio de Janeiro from March 01, 2020 to July 31, 2020. The format of the data is displayed in Table 2. The column `Symptom onset date` denotes the symptom onset date reported by the patient, and `Outcome date` marks the date on which the person recovers or dies (respectively `recovered` or `death` in the column `Outcome`), thus ceasing to be an active case. The column `Outcome date` was incomplete having about 3.7% of empty fields, so we were not able to use the recovery date as an input in our data fitting and missing death dates were imputed randomly according to the empirical distribution of the time difference between notification and evolution dates. Finally, we normalize the data by the size of the population in Rio de Janeiro, which we assume to be 6.7 millions (IBGE, 2021).

**Table 2:** Extract of the data: notification date, symptom onset date, neighborhood of residence, sex, age group, outcome, outcome date, ethnicity.

| Notification date | Symptom onset date | Neighborhood | Sex | Age group | Outcome | Outcome date | Ethnicity |
| --- | --- | --- | --- | --- | --- | --- | --- |
| 05/18/20 | 05/03/20 | PACIENCIA | M | 50 - 59 | death | 05/17/20 | Black |
| 04/25/20 | 04/02/20 | BARRA DA TIJUCA | M | 80 - 89 | death | 05/01/20 | White |
| 05/06/20 | 05/06/20 | CACHAMBI | M | 70 - 79 | death | 05/07/20 | Ignored |
| 06/12/20 | 06/02/20 | BARRA DA TIJUCA | M | 70 - 79 | death | 06/12/20 | White |
| 06/13/20 | 04/26/20 | MARECHAL HERMES | M | 60 - 69 | death | 06/16/20 | Ignored |

#### 2.2.1. Smoothed and cumulative curves

There is a weekly seasonality in confirmed cases and deaths in the outbreak with a negative deviation on weekends. This variation affects the model since it has no seasonal adjustment. For this reason, the data is smoothed using a centered *moving average* with 7 days. Thus, given a set of initial data {*x*_*s*_}_1≤*s*≤*n*_ of confirmed positive cases or deaths, we replace it with

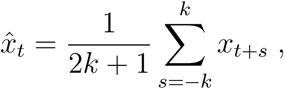

where 2*k* + 1 = 7. Although this choice may seem arbitrary, it is directly related to the weekly periodicity of the data. Figures 2 and 3 display daily new cases and deaths, respectively. In particular, in Figure 2 we can observe that the first wave ends at the end of July, which justifies our choice to set the horizon of the data fitting on July 31st. Moreover, we calculate the curves of cumulative positive cases and deaths by evaluating

**Figure 2:**
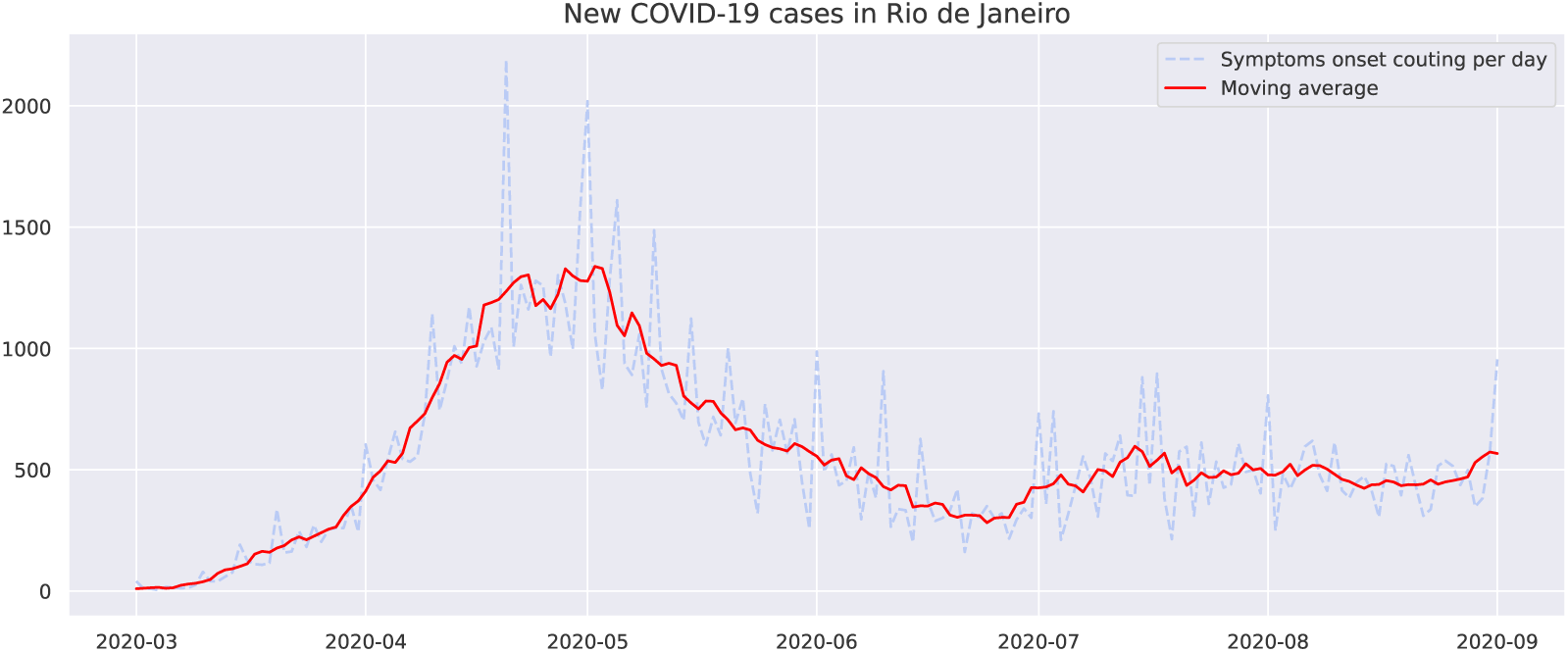
Daily new cases considering the self-reported symptoms onset in Rio de Janeiro between March and September of 2020. The 7-days moving average is displayed in red.

**Figure 3:**
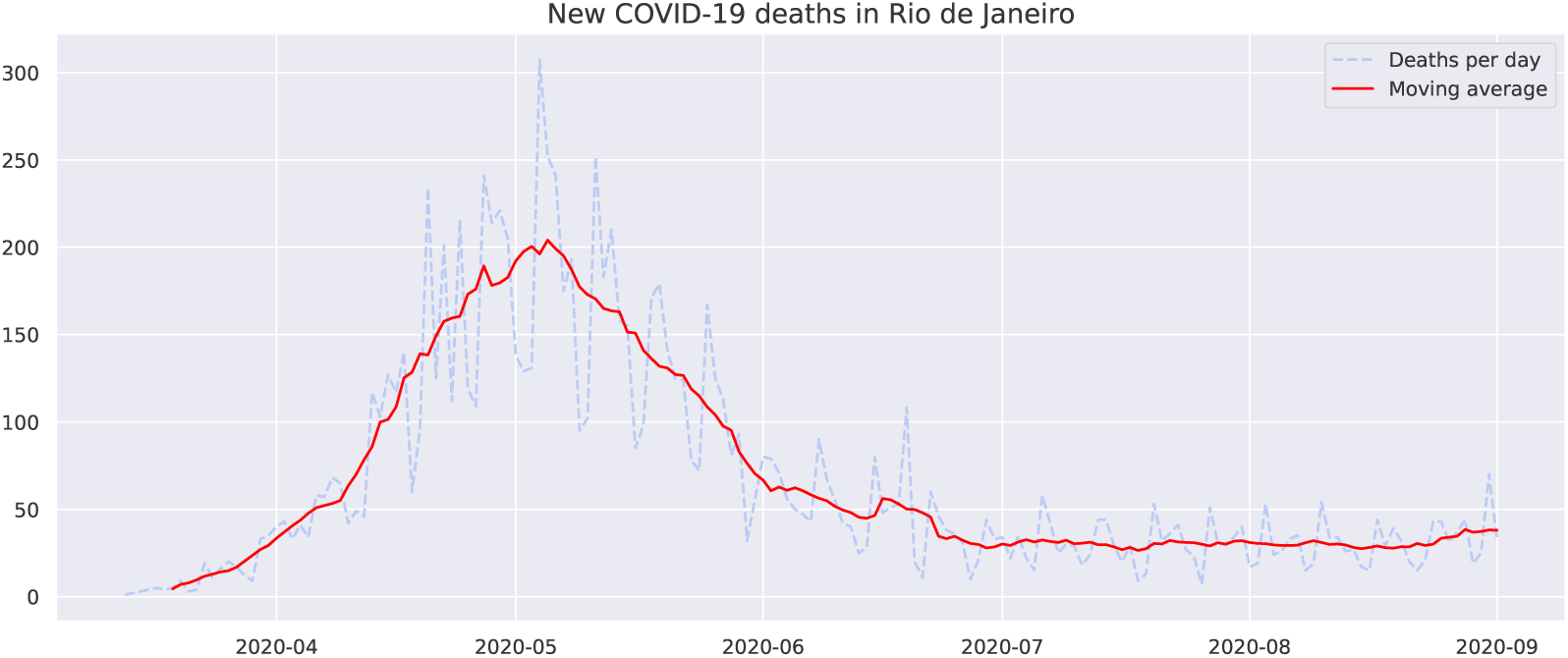
Daily new COVID-19 related deaths in Rio de Janeiro between March and September of 2020. The 7-days moving average is displayed in red.

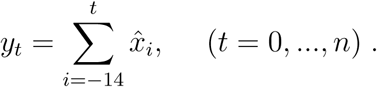

The curves of cumulative cases, considering the self-reported symptoms onset, and cumulative deaths will be compared to the compartments *T* and *D* from model (1).

### 2.3. Choice and modeling of parameters

We split the set of parameters between a first group, consisting of epidemiological constants *τ, σ, γ*_1_, *γ*_2_, and *δ*, whose values we retrieve from the literature (see Table 3), and a second group of parameters *β*(*t*), *α, ρ*, and *µ*(*t*) estimated from the data. The model used to represent the effective contact rate *β*(*t*) and the mortality *µ*(*t*) is described in Section 2.3.2. We start by estimating the testing rate *ρ*.

**Table 3:** Value of parameters retrieved from the literature.

| Par. | Value | Reference |
| --- | --- | --- |
| $\omega^{-1}$ | 5.74 days | Rai et al. (2021) |
| $\tau^{-1}$ | 3.69 days | Li et al. (2020) |
| $\sigma^{-1}$ | $\omega^{-1} - \tau^{-1}$ | Li et al. (2020) |
| $\gamma_1^{-1}$ | 7.5 days | Byrne et al. (2020) |
| $\gamma_2^{-1}$ | 13.4 days | Byrne et al. (2020) |
| $\delta$ | 0.01 | Kucirka et al. (2020) |

#### 2.3.1. Estimation of ρ

We consider the testing data from the state of Rio de Janeiro (IBGE, 2020), summarized in Table 4: about 2% of the population is tested each month, with a positive rate of about 20% among the tests done. We thus evaluate a daily testing of about 0.013% of the population, that is, we shall choose *ρ* ≤ 1.3 · 10^−4^. In this range of values, the impact of the choice of *ρ* on *α* is negligible, as we verify in Table 6. For this reason, we assign to *ρ* the value of *ρ* = 10^−5^. Unfortunately, the data do not allow to discern the tests due to symptom onset from the testing of asymptomatic cases, due to the effectiveness of the trace, track, and test strategy.

**Table 4:** Percentage of cumulative total and positive tests among the population.

|  | July | August | September | October |
| --- | --- | --- | --- | --- |
| Percentage (%) | 6.8 | 8.6 | 10.2 | 11.9 |
| Positive rate (%) | 1.2 | 1.5 | 1.9 | 2.4 |

#### 2.3.2. Representation of β and µ

The parameter *β* in model (1) varies in time according to the different public policies in place in Rio de Janeiro in different periods (Official Journal of the State of Rio de Janeiro, 2020b; Estadão, 2020), and according to the compliance of the population with these measures. For this reason, we approximate *β* by *B-splines* in the form

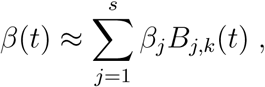

where *β*_*j*_ are the coefficients to be estimated and 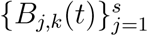 is the basis of functions of order *k* (De Boor, 1978). A similar representation is used for the mortality rate *µ*(*t*),

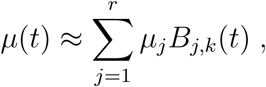

which has to be understood as relative to the number of confirmed deaths. This reflects the fact that the relative mortality may vary in periods of dis-tress for the health system and lack of testing kits. We therefore define the vector of *s* + *r* + 1 parameters to be estimated

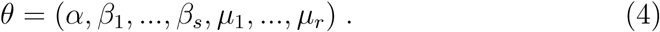

We assume that the *knots*, i.e. the time points where the polynomials connect, are equally spaced. The number of knots is equal to the number of parameters (*s* for *β* and *r* for *µ*) plus the order of the B-spline (*k*) plus 1.

To determine the order *k* and the number of coefficients *s, r*, for the B-spline approximation, we use the *Akaike Information Criterion (AIC)* (Liang et al., 2010), which under the normality hypothesis of the errors is given by the formula

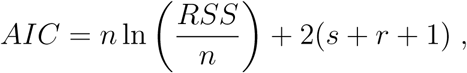

such that *RSS* is the sum of squares of the model’s residuals. For the numerical experiments in Section 4, we compare models with 3 and 4 coefficients, as models with less than 3 coefficients are unable to capture temporal variations over the timeframe of the outbreak, while models with *s* and *r* greater than 4 are computationally difficult to handle.

### 2.4. Parameter estimation

For the parameter estimation, we follow the methods applied in (Cao et al., 2012; Liang et al., 2010; Ramsay et al., 2007). The first 15 days of March were used to estimate the initial conditions (see Section 2.4.1) and thus set up the model starting from March 16th, when the first containment measures were imposed (Official Journal of the State of Rio de Janeiro, 2020a). Denoting by 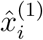 and 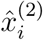 the daily new cases and deaths, respectively, on the *i*-th day, where *i* = 0 and *i* = *n* indicate March 16th and July 31th, respectively, we assume that

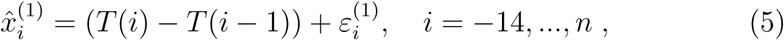

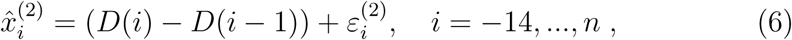

where the sequences 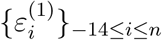 and 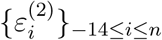 are independent and normally distributed random variables with unknown variances 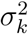, *k* = 1,2. Summing up we get

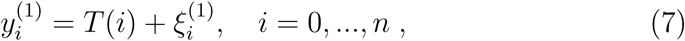

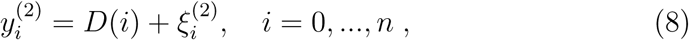

for the cumulative quantities, where 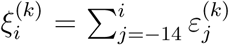 for *k* = 1, 2. Therefore, for any *i* ≤ *j* ≤ *n* and *k* = 1, 2, the *covariance matrix* is defined by

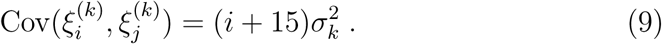

At this point, we introduce the notations 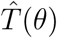 and 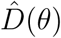 for the numerical approximations of the functions *T* and *D*, obtained by integrating equations (2) and (1), respectively, through the Runge-Kutta method and corresponding to the parameter vector *θ* introduced in (4).

#### 2.4.1. Initial values estimation

Considering *S* ≈ 1 in the first 15 days of the epidemic, and focusing on the compartments *E, I, A*, and *T*, the system (1) reduces to

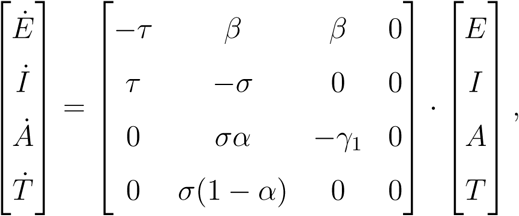

whose solution is a linear combination of exponential functions. The daily testing of asymptomatic individuals at the beginning of the pandemic is assumed to be negligible, thus *ρ* ≈ 0, while the parameters *γ*_1_, *τ*, and *σ* are taken from the existing literature (see Table 3). The parameters *α* and *β* are fixed in this period. The value *T* (−14) corresponds to the confirmed cases on March 2 and *T* (−15) is set to zero. Therefore, the parameters to be estimated in this period are reduced to *θ*_0_ = (*α, β, E*(−14), *I*(−14), *A*(−14)). Considering 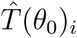 the approximation for *T* as function of *θ*_0_ at day *i*, we aim to minimize the expression

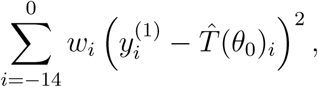

where the weights 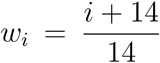 are chosen to give less importance to the initial days. With these estimates, we obtain the values (*E*(0), *I*(0), *A*(0)). Since *R*(−14) = 0, we get that *Q*(−14) = *T* (−14), and thus we can integrate the curves *Q* and *R* to obtain the values *Q*(0) and *R*(0). Finally, we deduce *S*(0) = 1 − *E*(0) − *I*(0) − *A*(0) − *Q*(0) − *R*(0).

##### Remark 2.1.

*In order to analyze the sensitivity of the curves* 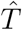 *and* 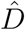 *with respect to the initial guess of the parameter θ*_0_, *we perform a series of random realizations and compare the final values* 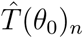 *and* 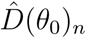 *for all guesses*.

#### 2.4.2. Curve fitting

We use the *weighted least squares* method to estimate the unknown parameters by the data of daily confirmed cases and deaths given by equations (7)-(8). This approach is based on the solution of a constrained non-linear minimization problem. We consider the following objective functional

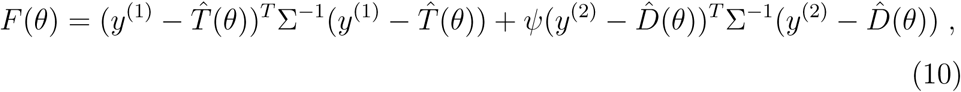

where *M* ^*T*^ denotes the transpose matrix of 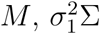 and 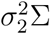 are the covariance matrices given by equations (9), and *ψ* is a weight proportional to the ratio of the variances 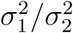. We solve the minimization problem by means of the L-BFGS-B algorithm (Byrd et al., 1995), which combines the gradient projection method and the BFGS algorithm with optimized use of computational memory, implemented in the SciPy library in Python (Virtanen et al., 2020).

### 2.5. Uncertainty quantification over the parameters

Uncertainty is intrinsic in the parameters’ estimation, owing to a combination of different factors, such as the natural variability of the data, the measurement errors of the data collection, and the biases of the estimation method. In order to tackle this problem, we construct confidence intervals for each unknown parameter. We rely on the *Bootstrap method* (Efron and Tibshirani, 1986), which involves a constructive approach based on the data. Starting from a series *Y*, it generates replicated data 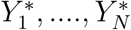 and performs the estimations for each one. The confidence interval for the value of interest is then given by the corresponding percentile of the *N* replicated samples (Joshi et al., 2006).

As a consequence of the error structure presented in equation (5), we generate each replicated curve *T* ^(*B*)^ with the initial value 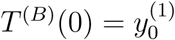 and satisfying *T* ^(*B*)^(*i* + 1) = *T* ^(*B*)^(*i*) + *ε*_*i*+1_ for any *i* ≥ 1, where 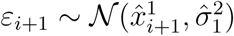 and 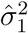 (see Appendix A) is an estimate for 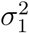 An analogous construction is carried out for equation (6). In the estimation process, in order to avoid local minima, we randomize the initial guess of the optimization algorithm: for every *j* = 1, …, *r* + *s* + 1, we select initial guesses 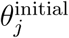 randomly chosen with uniform distribution in a prescribed range (*l*_*j*_, *u*_*j*_), that represents the interval of admissible values for that parameter. After *m* iterations of this process, we pick the best solution minimizing the objective functional (10).

## 3. Theory

### 3.1. Identificability

From a structural-theoretical point of view, we are interested in analyzing the *identifiability* of model (1). In general, this analysis identifies whether unknown parameters can be estimated in a unique way from the available measurements on the system (Audoly et al., 2001; Saccomani and Thomaseth, 2019). There are two conceptually different ways to develop this analysis: the structural approach (a priori) and the practical approach (a posteriori). The former is a theoretical property that resides in the structure of the model itself: the parameters are *structurally identifiable* if they can be (globally) uniquely identified from the available measurements; they are called *locally structurally identifiable* if they are structurally identifiable within a neighbourhood of the solution. On the other hand, the practical approach is based on the outcome of the data fitting, and it is evaluated by means of the *correlation matrix* of the parameters.

### 3.2. Structural identifiability

Consider a dynamical system

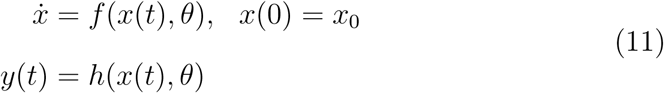

where *x*(*t*) ∈ ℝ^*n*^, *y*(*t*) ∈ ℝ^*m*^ and *f* and *h* are rational functions of the variable *x* and *θ* ∈ Θ ⊂ ℝ^*p*^, Θ being the set of admissible values of the parameter *θ*. The variable *y* is the output of the system, that is the observable component. In the model, *y* is the number of confirmed cases and deaths. When *x*(0) = *x*_0_, the measurement with initial value *x*_0_ is 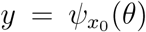. The structural identifiability (Ljung and Glad, 1994) of system (11) at a fixed *θ*\* ∈ Θ is related to the number of solutions of the equation

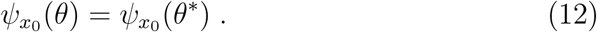

The system (11) is *globally identifiable* at *θ* * if equation (12) has a unique solution *θ* = *θ*\*, or, equivalently if the mapping 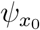 is invertible. System (11) is *locally identifiable* if 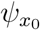 is invertible in a neighborhood of *θ*\*. In Appendix B we describe a computational method to verify structural identifiability using the DAISY software (Bellu et al., 2007).

### 3.3. Practical identifiability

The structural identifiability process developed in Section 3.2 is based on two hypotheses: the model structure is accurate and measurement errors are absent. These hypotheses are not valid in practice and, therefore, it is necessary to assess whether the parameters can be reliably and accurately estimated from noisy data (Miao et al., 2011). Let 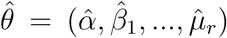 be the vector of parameters estimated from the data fitting, and 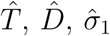 and 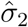.the model approximations for the confirmed cases, deaths, and their respective estimated variances, as introduced in Section 2.4. The correlation matrix quantifies the interdependence between model parameters, and can be computed as follows: starting from the *Fisher Information matrix (FIM)*

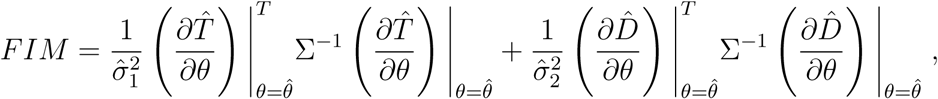

we compute the *covariance matrix C* as the inverse of *FIM*. Then, the element *r*_*ij*_, 1 ≤ *i, j* ≤ *r* + *s* + 1 of the *correlation matrix R* is defined as

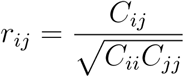

which measures the correlation between 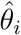 and 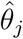 A value close to 1 indicates that the parameters are strongly interconnected, and that each of them varies according to the other.

## 4. Results

We first set the initial conditions according to Section 2.4.1 and we approximate the curve of the first two weeks of the outbreak, as represented in Figure 4. Following Remark 2.1, the fitting with respect to the initial guess for *θ*_0_ is robust: the final values on July 31st range from 0.0127 to 0.0131. As summarized in Table 5, the best fitting over the outbreak period March-July 2020 is obtained by choosing an approximation with 4 coefficients for the two parameters *β* and *µ*, and B-spline of order 2 for *β* and order 1 for *µ*.

**Table 5:**
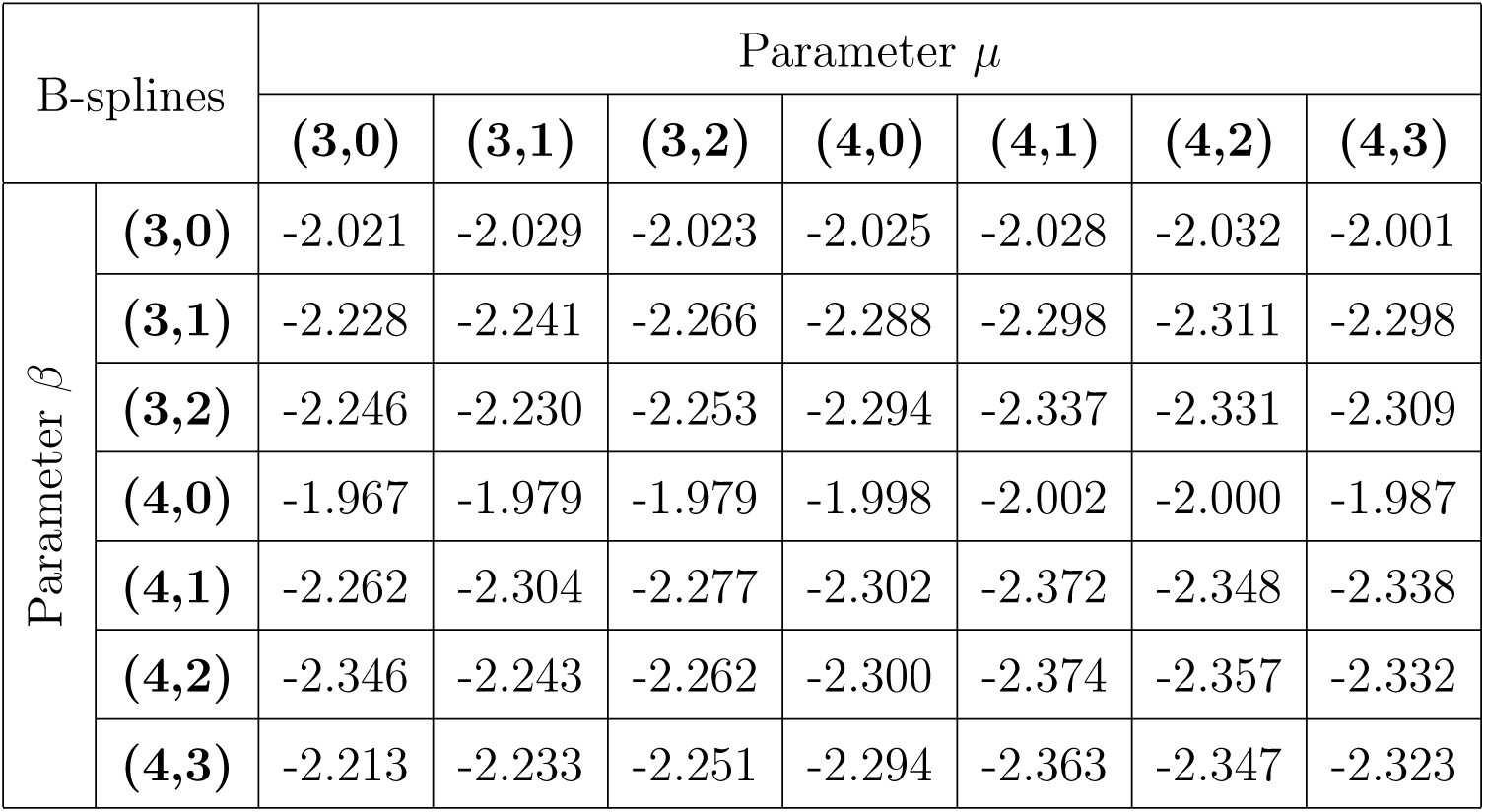
Model selection according to AIC (10^3^ scale): (*r, k*) represents the number of coefficients and the order of the B-spline.

**Figure 4:**
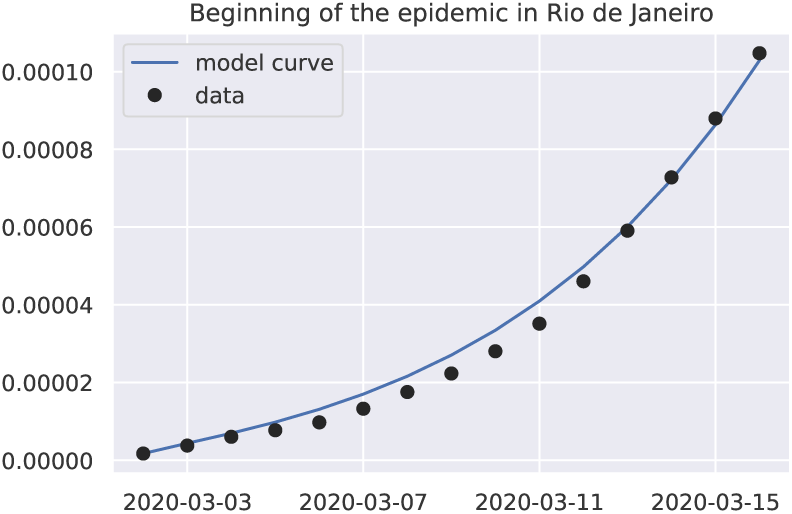
Fitting the curve of cumulative cases at the beginning of the outbreak

We fit the available data using the L-BFGS-B algorithm explained in Section 2.4.2. Figure 5 shows a very good matching between the available data and the fitting curves. To analyze the robustness of the estimate of *α* with respect to the epidemiological parameters, we perform the following analysis: choosing a grid of values based on the confidence interval of each parameter, we estimate the parameters for each possible combination and we determine the variation in the estimated value of *α*, as reported in Table 6.

**Figure 5:**
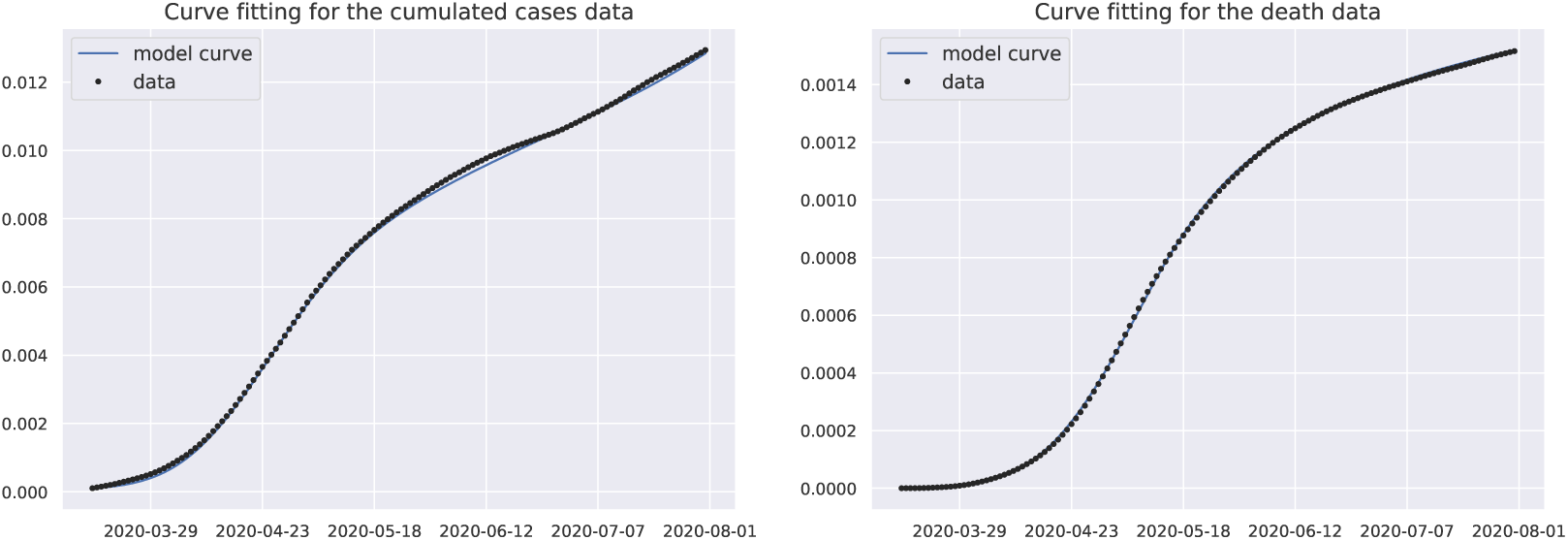
Fitting the curves of cumulative cases and deaths

**Table 6:** Range of values of *α* corresponding to admissible intervals of the parameters according to the references of Table 3

| Parameters | Interval | Range of values of $\alpha$ |
| --- | --- | --- |
| $\tau^{-1}$ | $[2, 4]$ | $[0.897, 0.902]$ |
| $\sigma^{-1}$ | $[2, 4.5]$ | $[0.897, 0.9]$ |
| $\rho$ | $[0, 10^{-4}]$ | $[0.898, 0.899]$ |
| $\gamma_1^{-1}$ | $[6.5, 9.5]$ | $[0.894, 0.903]$ |
| $\gamma_2^{-1}$ | $[11, 16]$ | $[0.896, 0.898]$ |

We also verify whether the model residuals approximate the errors, which are assumed to have a Gaussian distribution. The histograms of the residuals and the *Q-Q plots* (comparison of the quartiles of the Gaussian distribution with the quartiles of the sampling distribution of the residuals) are shown in Figure 6. In addition to this visual analysis, we can also apply statistical tests to verify correlation and normality: the *Ljung-Box test* (Ljung and Box, 1978) returns a p-value 0 which indicates that the residuals are not uncorrelated; the *Jarque-Bera test* (Jarque and Bera, 1980) does not reject the null hypothesis at the 5% level, which is an evidence supporting the normality of the residuals.

**Figure 6:**
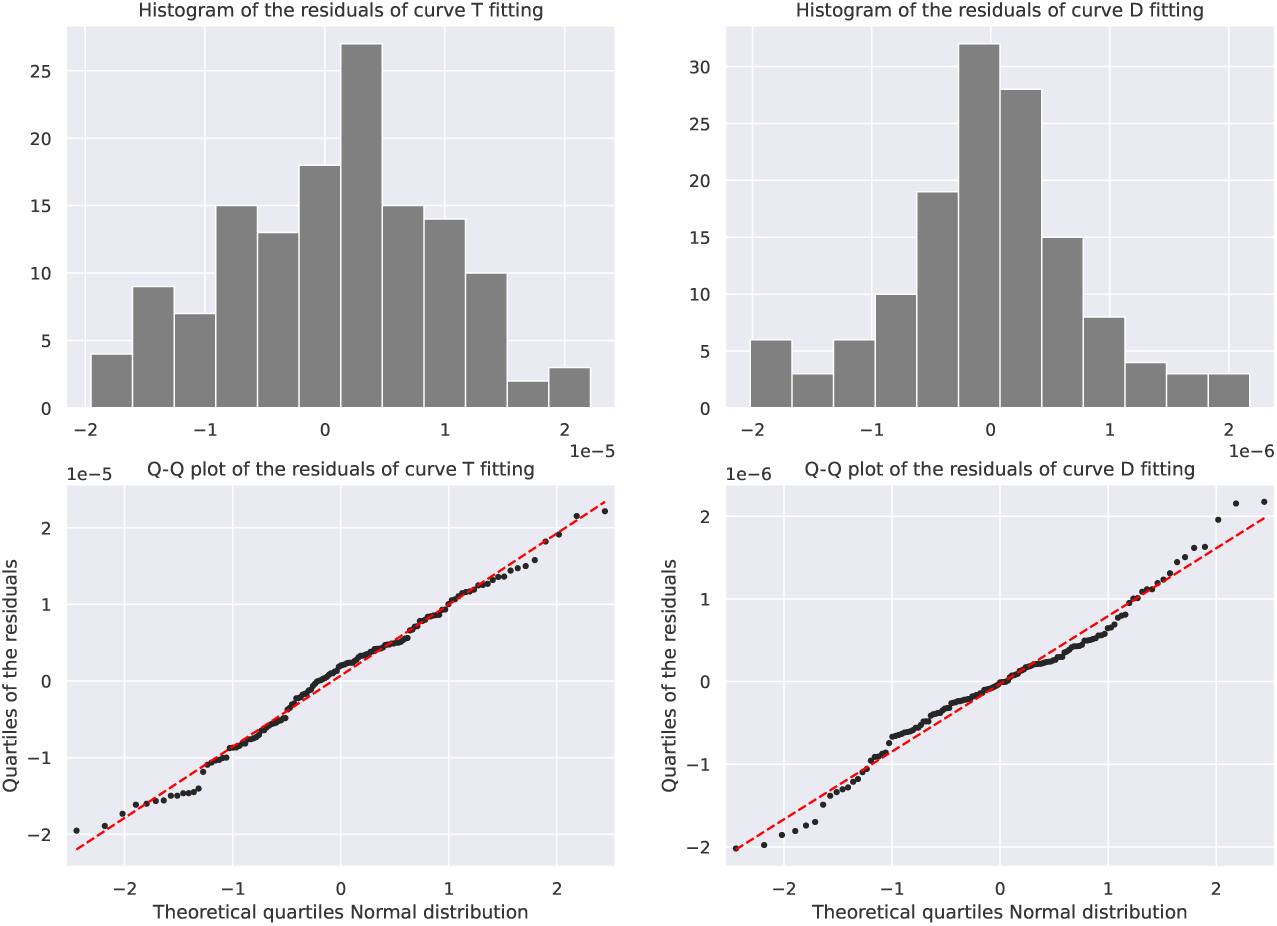
Graphical analysis of the model’s residuals with histogram e Q-Q plot.

Another important outcome of the study is the estimate of the effective (time-varying) reproductive number ℛ_*t*_. Figure 7 describes the evolution of ℛ_*t*_ in the period under consideration.

**Figure 7:**
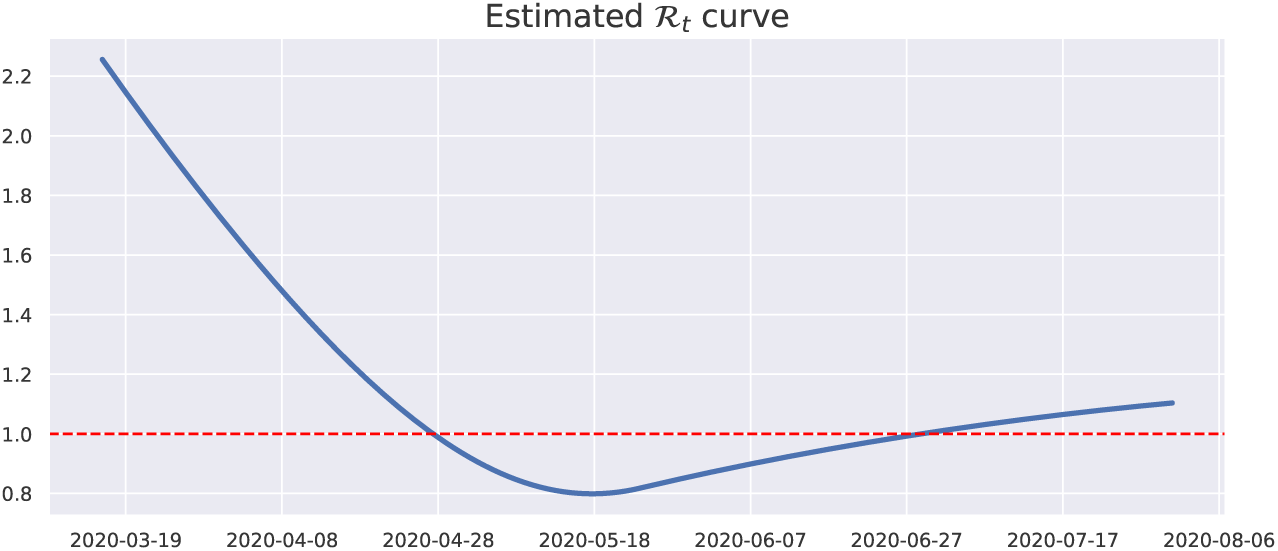
Effective (time-varying) reproduction number ℛ_*t*_. The dashed red line indicates the threshold value ℛ_*t*_ = 1

The correlation matrix among the estimated parameters (Section 3.3) is depicted in Figure 8. It is worth noticing that the parameter *α* and the second coefficient of the B-spline of *β* have a strong inverse correlation.

**Figure 8:**
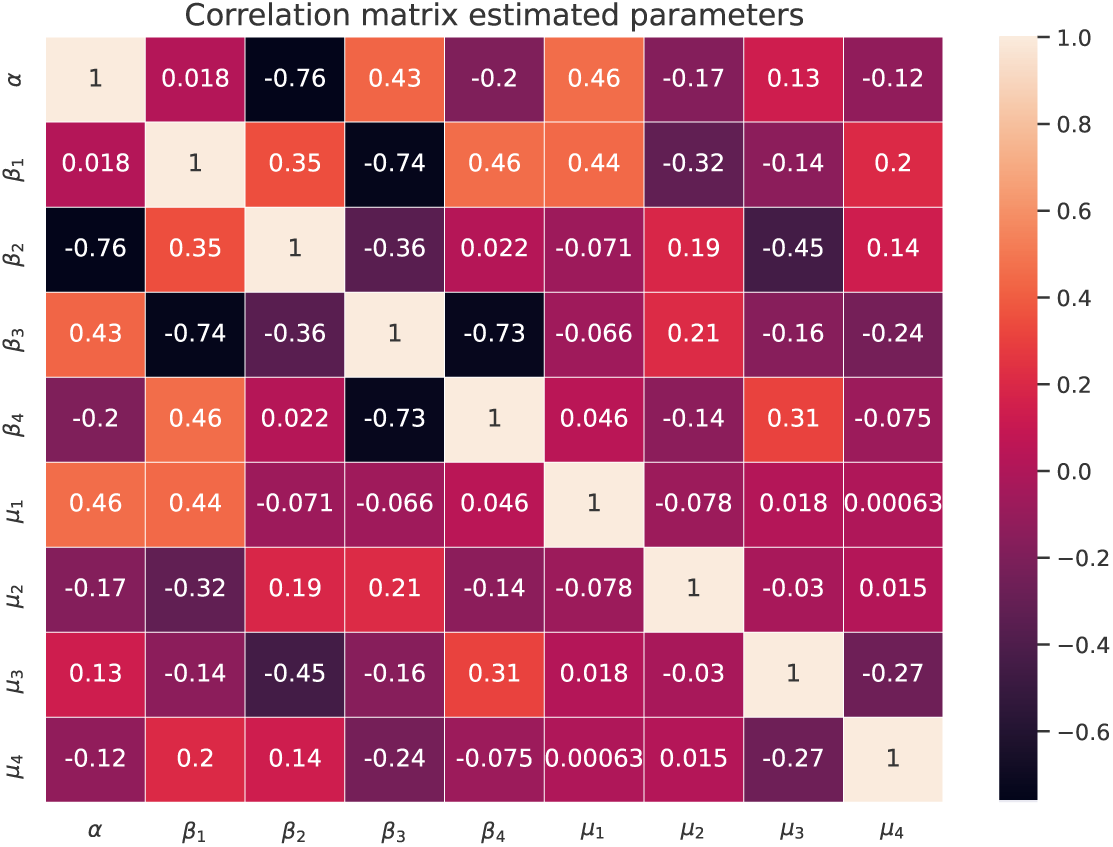
Correlation matrix of the estimated model’s parameters.

Relying on the Bootstrap method from Section 2.5 we derive confidence intervals for the parameters of the fitting: after *N* = 500 simulations with *m* = 10, we conclude that the 95% confidence interval for the unreported rate *α* is (0.849, 0.931). Figure 9 displays the confidence interval of ℛ_*t*_ over time. Figure 10 provides a scatter plot and histogram visualization for the estimated correlations in Figure 8. Finally, Table 7 summarizes the estimates for all parameters.

**Table 7:** Estimates of the median and confidence intervals for each parameter.

| Parameter | Median | Confidence interval |
| --- | --- | --- |
| $\alpha$ | 0.903 | [0.849, 0.93] |
| $\beta_1$ | 0.144 | [0.142, 0.147] |
| $\beta_2$ | 0.073 | [0.069, 0.079] |
| $\beta_3$ | 0.101 | [0.098, 0.104] |
| $\beta_4$ | 0.126 | [0.123, 0.132] |
| $\mu_1$ | 0.0101 | [0.0092, 0.011] |
| $\mu_2$ | 0.0159 | [0.0154, 0.0164] |
| $\mu_3$ | 0.0103 | [0.0098, 0.0108] |
| $\mu_4$ | $7.03 \cdot 10^{-3}$ | $[6.73 \cdot 10^{-3}, 7.38 \cdot 10^{-3}]$ |

**Figure 9:**
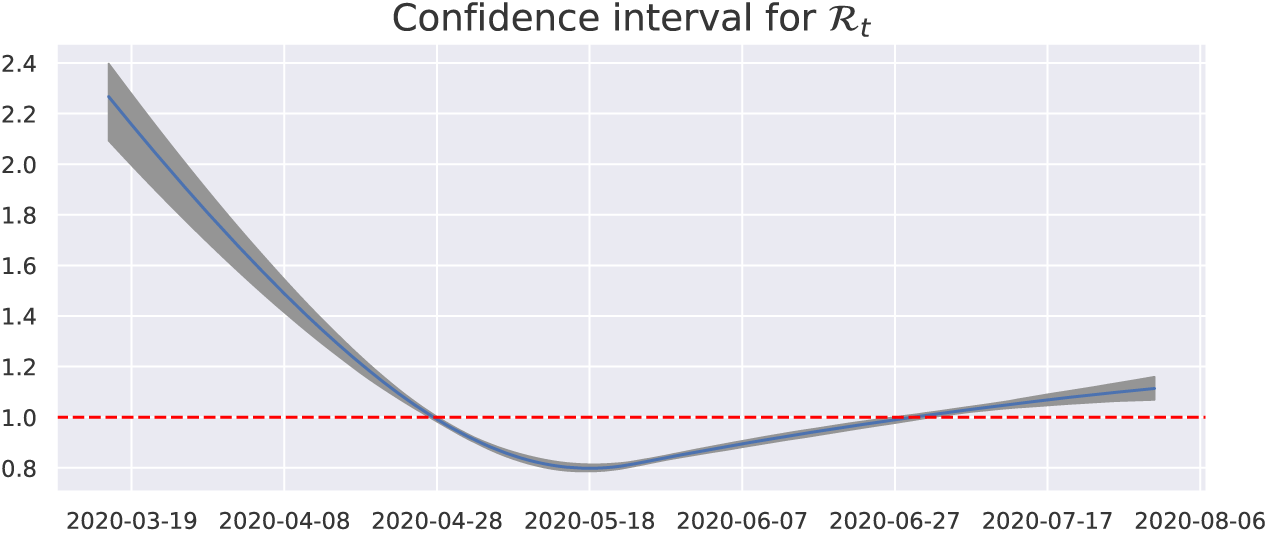
The blue curve is the median of the estimated curves and the gray represents the confidence interval for each time. The dashed red line is the threshold 1 for ℛ_*t*_.

**Figure 10:**
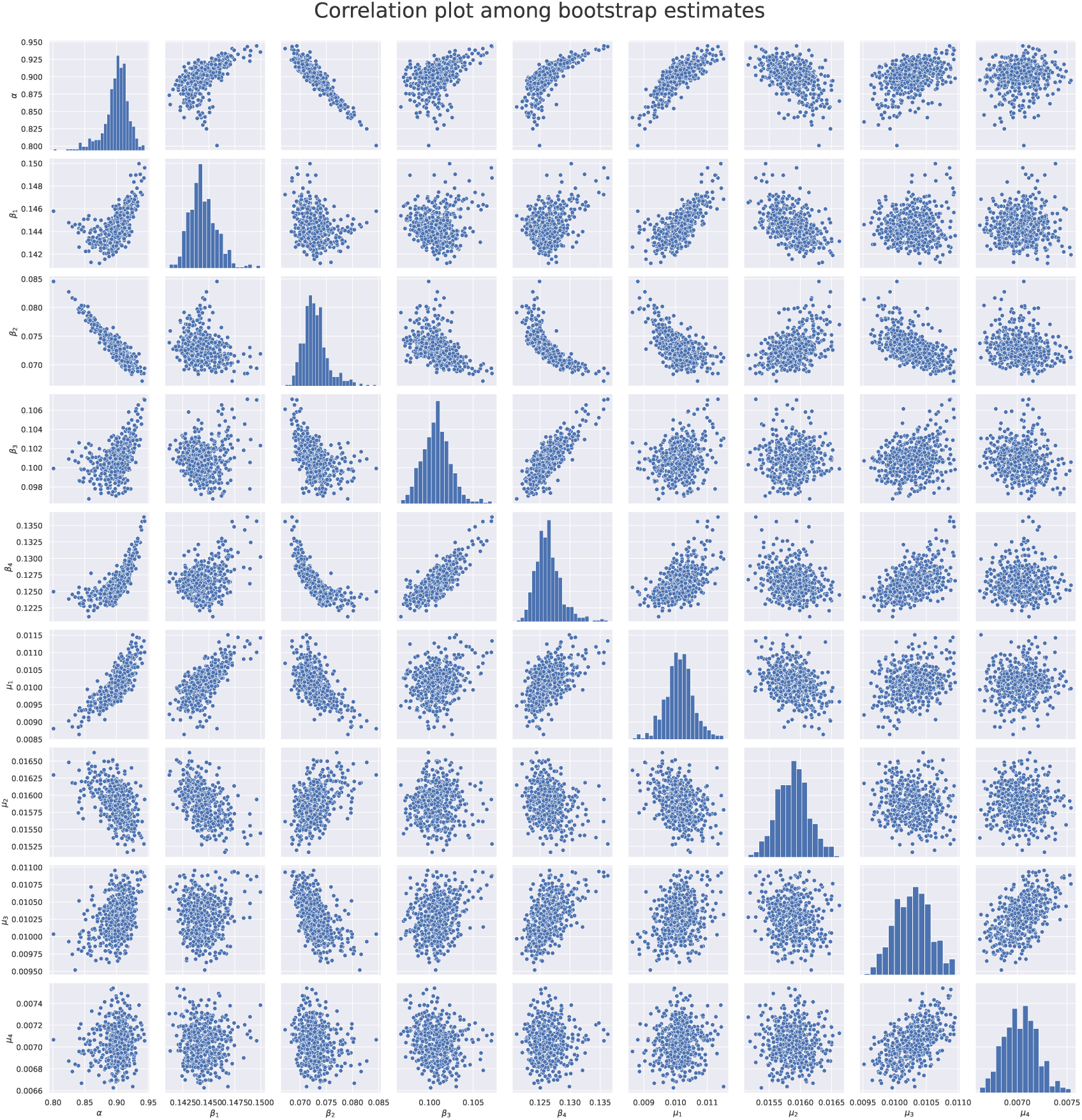
Scatter plots for each pair of two parameters, and their corresponding histogram.

## 5. Discussion

The range of values for the unreported rate *α* is in line with the results of other works on this issue: in Rio de Janeiro, (Prado et al., 2020) estimated the notification at 7.2%; at a Brazilian national level, (Canzian, 2020; Prado, 2021; Portal COVID-19 Brasil, 2021) estimated the notification of cases in the range [7.8%, 8.1%].

It is also interesting to compare the estimates of ℛ_*t*_ over time: in (Mellan et al., 2020), the point estimate for May 9, 2020 from the state of Rio de Janeiro was 1.1 with 95%-confidence interval [0.9, 1.3], which includes our range of values. The curve estimated by (Observatório COVID-19 BR, 2021) for the city of Rio de Janeiro has a very similar trend as in Figure 7: at the beginning of May, the ℛ_*t*_ has an estimated value smaller than 1 and it grows again until it is greater than 1 at the end of July. These values corroborate our fitting.

The correlation between *α* and *β*_2_ represents a limitation in terms of identifiability of the system. In order to achieve a structural identifiability result it is necessary to know the recovered curve (see Appendix B), whereas for the practical identifiability it would be useful to approximate the transmission and mortality functions with a different model.

## 6. Conclusions

In this work, we combine tools from the analysis of differential equations, statistics, and optimization to estimate the unreported rate of COVID-19 in the city of Rio de Janeiro during the first outbreak in March-July 2020. We determine that the rate of unreported positive cases is about 90%, with a confidence interval between 85% and 93%. This means that every case reported by the health system corresponds to about 9 – 10 cases that were not detected. This estimation shall be considered as a statistical approximation of the unreported rate: in addition to the lack of testing kits during the period under evaluation, other delays and errors at all stages of the notification process may generate bad inferences. Data analysis techniques have been deployed to deal with the low quality of data.

Concerning the modeling chosen for the fitting, it is of great importance to underline that the initialization of the parameters has little influence on the final estimation of *α*, supporting the robustness of our analysis. However, the fact that the residuals of the estimation are normally distributed but correlated needs more attention and will be addressed in future works.

Finally, the outcome of this work provides a description of the evolution in time of the disease reproductive number. This estimation provides a useful tool to determine periods of growth or decrease of the epidemic force at a geo-localized level, and thus to inform policymakers in their decision process to limit the spread of the virus and its consequences on the population.

## Data Availability

All data produced in the present study are available upon reasonable request to the authors

## Appendix A

### Variance estimate

For the variances 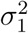 and 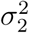, we follow the estimates from (Seber and Wild, 2005, p. 21-28) given by

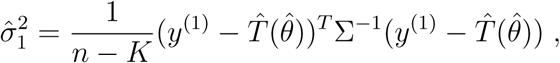

where *K* = *r* + *s* + 1 and *n* is the number of data points. In a similar way,

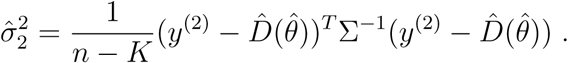

## Appendix B

### DAISY

DAISY - Differential Algebra for Identifiability of SYstems - is a software based on the programming language *Reduce*, specific to the problem of identifiability in dynamical systems. DAISY requires certain specific conditions on the system, that allow to apply algebraic methods to the model under consideration. In order to rewrite our system (1) in terms of the DAISY algorithm, we treat all model parameters’ in Section 2.1 as constant in time. If we assume to know the curve *R* of recovered cases, we verify that model (1) is globally identifiable. However, the knowledge of *R* may sometimes be only partially available, as discussed in Section 2.2. Without access to these data, DAISY has not been able to certify the structural identifiability of the system.

## Acknowledgements

The authors wish to thank Luiz Max Carvalho (FGV EMAp) and Marcelo Fernandes (FGV EESP) for fruitful discussions about several statistical methods applied in this research.

The first and third authors were supported by FAPERJ and CNPq, Brazil. The second author acknowledges the funding of the Natural Sciences and Engineering Research Council of Canada (NSERC). The third author thanks the Center for the Development of Mathematics and Science (FGV CDMC) for their support.

## Notes

### Competing Interest Statement

The authors have declared no competing interest.

